# Triaging patients in the outbreak of the 2019 novel coronavirus

**DOI:** 10.1101/2020.03.13.20035212

**Authors:** Guoqing Huang, Weiqian Zeng, Wenbo Wang, Yanmin Song, Xiaoye Mo, Jia Li, Ping Wu, Ruolong Wang, Fangyi Zhou, Jing Wu, Bin Yi, Zeng Xiong, Lu Zhou, Fanqi Wang, Yangjing Tian, Wenbao Hu, Xia Xu, Ruonan Zhai, Kai Yuan, Xiangmin Li, Xinjian Qiu, Jian Qiu, Aimin Wang

## Abstract

In the end of 2019, the epidemic of a new coronavirus (SARS-CoV-2) occurred in Wuhan and spread rapidly. Changsha, a city located south to the epicenter, was soon impacted. To control the transmission of the coronavirus and avoid nosocomial infection, triage procedures based on epidemiology were implemented in a local hospital of the city. This retrospective study analyzed the data collected during the triage period and found that COVID-19 patients were enriched seven folds into the Section A designated for rapid detection and quarantine. On the other side, roughly triple amounts of visits were received at the Section B for patients without obvious epidemiological history. Eight COVID-19 cases were spotted out of 247 suspected patients. More than 50% of the suspected patients were submitted to multiple rounds of nucleic acid analysis for SARS-CoV-2 infection. Of the 239 patients who were diagnosed as negative of the virus infection,188 were successfully revisited and none was reported as a COVID-19 case. Of the eight COVID-19 patients, three were confirmed only after multiple rounds of nucleic acid analysis. Besides comorbidities, delayed sharing of epidemiological history added another layer of complexity to the diagnosis in practice. While SARS-CoV-2 epidemic is being alerted in many countries, our report will be helpful to other colleagues in rapid identification of COVID-19 cases and controlling the transmission of the disease.

## INTRODUCTION

The current epidemic of Coronavirus Diseases 2019 (COVID-19) associated with the severe acute respiratory coronavirus 2 (SARS-CoV-2) occurred in Wuhan (Hubei province) in December 2019 and rapidly spread to other areas in China and more than 100 other countries.^1-4^ According to the World Health Organization (WHO), as of March 1, 2020, the cumulative confirmed cases of COVID-19 in China have reached 79 968 with 2873 deaths (fatality rate 3.6%).^5^ More than 60% of the confirmed cases were reported from Wuhan.^6^ Besides SARS-CoV and MERS-CoV, SARS-CoV-2 is the third coronavirus species from the genus *Betacoronavirus* that leads to major epidemics in 21^st^ century.^3^ To prevent the disease transmission, China has suspended all public transport in and out of Wuhan since January 23, 2020.^7^

Changsha, a city located 350 kilometers south of the epicenter, reported its first imported COVID-19 case on January 21 and the highest level of public health emergency response was declared two days later.^8^ The common symptoms of COVID-19 patients at illness onset include fever, cough, expectoration, headache, myalgia or fatigue, diarrhoea and haemoptysis.^1,9,10^ Some of these symptoms resemble much like other diseases including flu, which has high occurrence in winter. To avoid the transmission of SARS-CoV-2 within hospital, triage procedures for patients were implemented between January 28 and February 20 in a local hospital (Xiangya Hospital) of Changsha to facilitate the rapid detection and quarantine of COVID-19 patients. Here we describe the clinical practice of triaging patients in the epidemic of SARS-CoV-2, along with the clinical and laboratory characteristics of eight COVID-19 cases identified from more than 240 suspected individuals with various symptoms triaged to the section for patients without obvious epidemiological history.

## METHODS

### Triage and patients

This single-centre, retrospective, observational study was done at Xiangya Hospital (Changsha, China). Patients with fever, respiratory symptoms, myalgia, fatigue, or other symptoms possibly related to SARS-CoV-2 infection were received at the triage reception between January 28 and February 20 before being directed to the Section A or B based on epidemiological characteristics.^11^ Patients, who met one of the following conditions within 14 days before illness onset, were sorted to the Section A: (a) exposure to Hubei province or local communities with confirmed COVID-19 cases reported; (b) exposure to patients with similar symptoms from regions mentioned in (a); (c) exposure to known COVID-19 patients; (d) association with clustering occurrence (Figure 1A). Other patients were directed to the Section B.

**Figure 1.**
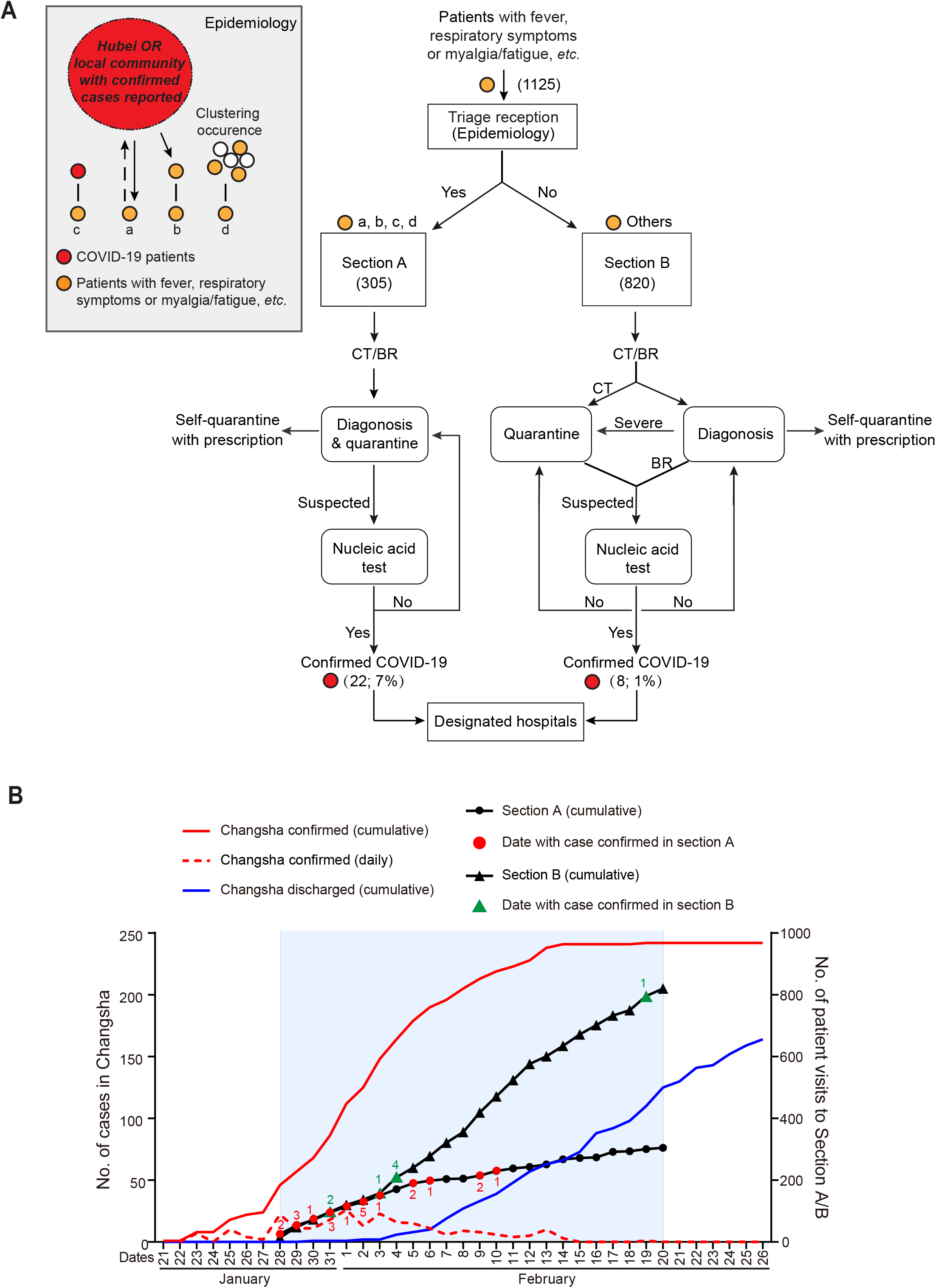
The triaging process in the local hospital. (A) The flowchart of triaging procedure. In total, 1125 visits were triaged to the Section A (305 visits) and B (820 visits). Suspected patients based on epidemiological history and CT/BR were analyzed for SARS-CoV-2 infection by real-time RT-PCR. 22 and 8 COVID-19 patients were identified in the Section A and B, respectively. The inset represents the epidemiological characteristics for triaging. See text for more details. BR: blood routine. (B) The epidemic situation of COVID-19 in Changsha. Confirmed and discharged cases of COVID-19 in Changsha are plotted in red and blue, respectively (left axis). The triaging period (January 28– February 20) is shaded. Cumulative visits to the Section A and B (right axis) are plotted in black circle and triangle, respectively (days with COVID-19 patients confirmed are marked in red and green with the number of patients indicated).

Besides symptoms, clinical and laboratory characteristics suggestive for SARS-CoV-2 infection are: (i) chest computed tomographic (CT) results with pneumonia features; (ii) normal or reduced leucocyte count or reduced lymphocyte count in early onset.^1,9,12,13^ At the Section A, patients having mild symptoms without both (i) and (ii) were recommended for home quarantine with prescription. Others were quarantined to take oropharyngeal swab (if not specified otherwise) for SARS-CoV-2 nucleic acid analysis by real-time reverse transcription polymerase chain reaction (RT-PCR). At the Section B, patients with suspected chest CT characteristics (i) were also quarantined for nucleic acid analysis. Others were further evaluated by doctor based on symptoms (fever, respiratory symptoms, myalgia/fatigue, *etc*.), comorbidities, vital signs and blood routine characteristics: patients with severe symptoms were quarantined for treatment and nucleic acid analysis and were transferred to relevant units for further treatment when diagnosed as negative for SARS-CoV-2 infection; patients having mild symptoms without (ii) were recommended for home quarantine with prescription, and the rest were submitted for nucleic acid analysis and were recommended for home quarantine with prescription if negative result of SARS-CoV-2 was obtained (the second nucleic acid test after 24 hours was recommended and performed based on patient’s availability) (Figure 1A). Self-quarantined patients were followed up by phone visiting. Identified COVID-19 patients were immediately transferred to designated hospitals for quarantine and treatment. All medical personnel working at both sections and the triage reception were equipped with appropriate protections.^14^ Suspected patients submitted for nucleic acid analysis at the Section B were enrolled in this study. The Ethics Commission of Xiangya Hospital approved this study (No. 202003031). Written informed consent was waived due to the rapid emergence of this infectious disease.

### Data collection

Blood routine, biochemical, radiological and microbiological data together with demographics, epidemiological characteristics, medical histories and vital signs (body temperature, heart rate, respiratory rate, blood pressure, blood oxygen saturation) of patients were collected from a local server. If data were missing from the records or clarification was needed, data were obtained by direct communication with patients, attending doctors, or other healthcare providers. All data were checked by two physicians (W. Wang and G. Huang). Patients with negative results of RT-PCR for SARS-CoV-2 infection were revisited by phone when applicable.

### Laboratory test

Clinical specimens for SARS-CoV-2 diagnostic test were obtained in accordance with clinical guidelines.^15^ Oropharyngeal and nasopharyngeal swabs were collected with synthetic fiber swabs, maintained in 2-3 mL viral-transport medium and stored between 2 °C and 8 °C until ready for test. RNA was extracted following the manufacture instruction (SANSURE). Laboratory confirmation of SARS-CoV-2 was performed using real-time RT-PCR kit following the manufacture instruction (SANSURE) on the ABI Q5 PCR machine. Analysis for influenza A/B virus was performed using antigen detection reagent (colloidal gold method). Routine bacterial examinations were also performed.

## RESULTS

In total, 1125 patients (visiting number) were received at the triage reception (January 28–February 20). Following the triage procedures, 305 visits were directed to the Section A, and 22 COVID-19 cases (7%, 22/305) were confirmed. In the first ten days of triage, confirmed cases were identified nearly every day (Figure 1B). On the other side, 820 visits were directed to the Section B, and eight cases (1%, 8/820) were confirmed (Table S1). Seven of them were spotted in the first ten days (Figure 1B). The implemented triage procedures effectively enriched COVID-19 patients into the Section A and reduced the possibility of transmitting the virus to other patients and medical stuff. During the triaging period, the number of reported COVID-19 patients in Changsha rapidly increased ten times (from 24 to 242) and reached plateau after February 14 (Figure 1B).

Of the 820 visits in the Section B, 239 individual patients were suspected, but laboratory evidence did not support for SARS-CoV-2 infection. 52% (124/239) of them did multiple rounds of nucleic acid test. 188 of 223 patients with contact information were successfully followed up by phone visiting a few days after the last nucleic acid test. None was reported as a COVID-19 case. 89% (167/188) of the patients were phone visited more than a week after their last nucleic acid test (Figure 2). 7% (13/188) of them were double checked by additional medical institutions.

**Figure 2.**
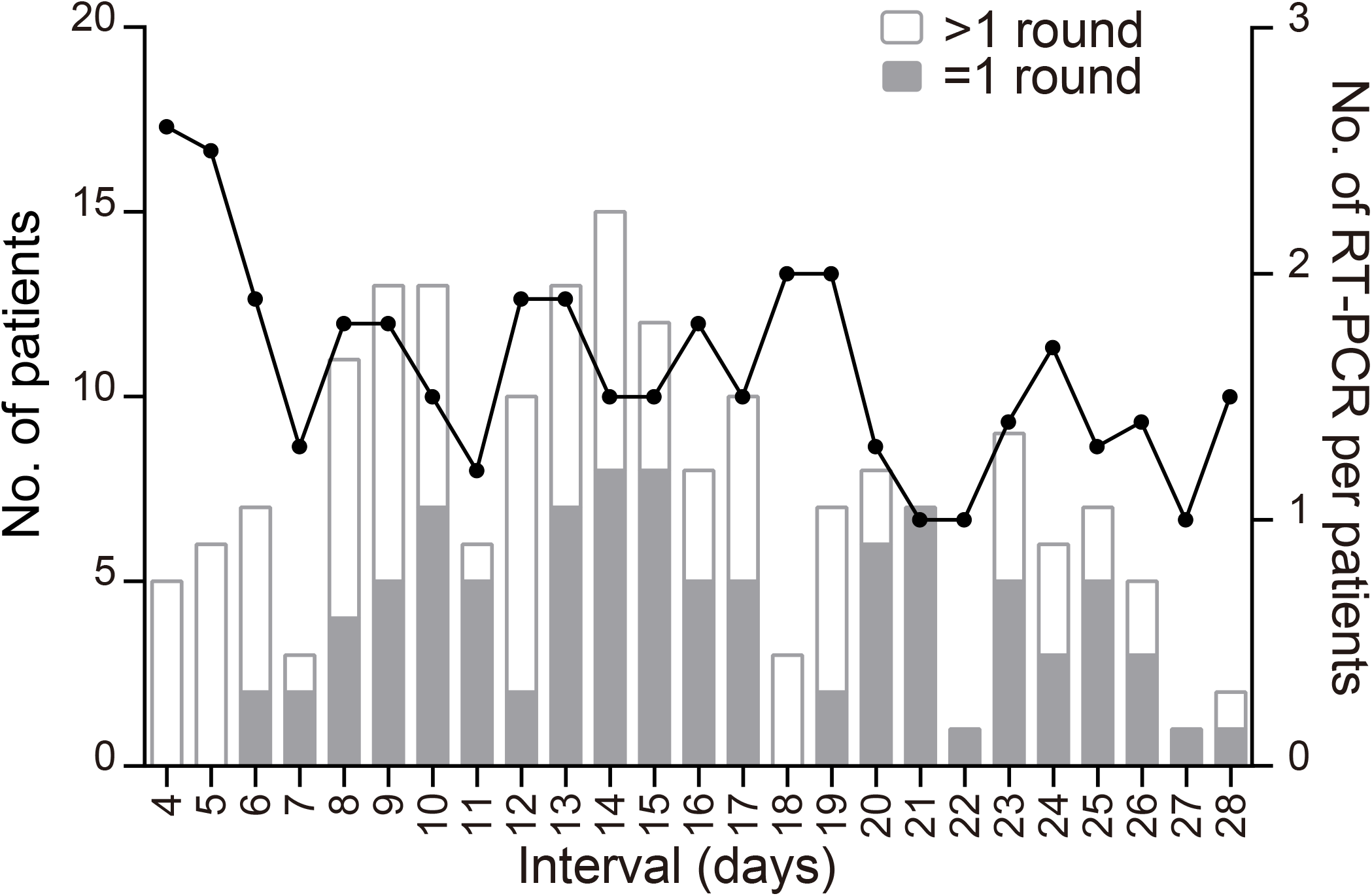
Follow-up of 188 patients excluded for SARS-CoV-2 infection by the Section B. The number of patients visited by phone (bar, left axis) and the average number of RT-PCR analysis per patient (black line, right axis) is plotted against the time interval between the phone visiting and the last RT-PCR analysis performed in the Section B. No patient was found to be a COVID-19 case after the last diagnosis in the Section B as negative for SARS-CoV-2 infection.

Of the eight COVID-19 patients identified by the Section B, five (cases 1–5) were confirmed with SARS-CoV-2 infection after one round of nucleic acid analysis (Figure 3 and Table S2). Cases 1 and 2 had no obvious epidemiological history. Cases 3, 4 and 5 (a familial cluster) arrived at the Section B initially without revealing both the contact history with people from Wuhan on a family event as well as the common symptoms among themselves. They were quarantined after the critical epidemiological information was given to the doctor during diagnostic inquiry and were soon confirmed to be positive for SARS-CoV-2 infection (Figure 3). Case 6 was evaluated as suspected COVID-19 case after arriving at the Section B (according to the chest CT images provided by another hospital) and was quarantined for further diagnosis. Nucleic acid analysis was performed on the first and the third day during quarantine. The second analysis supported for the virus infection (Figure 3).

**Figure 3.**
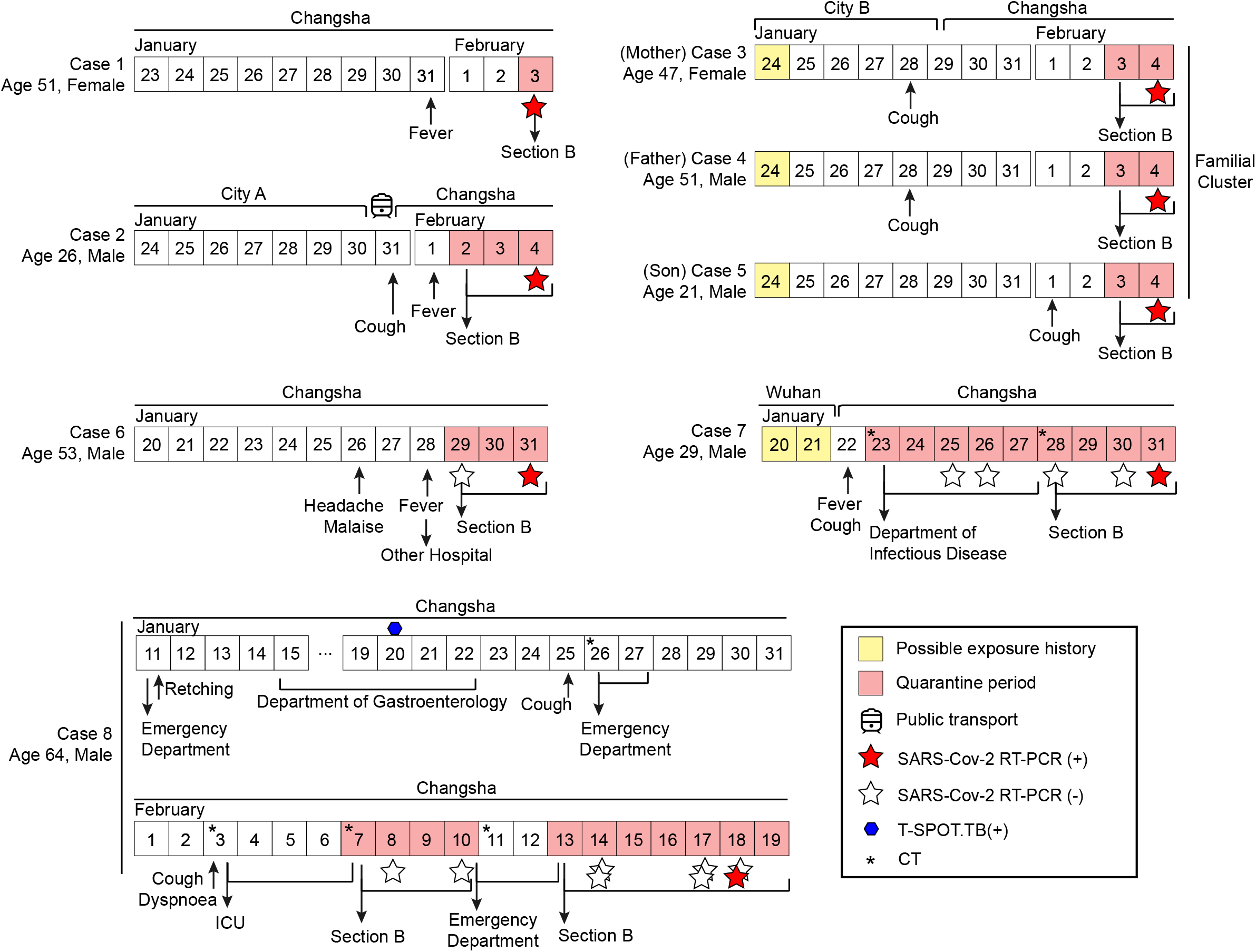
Timeline of illness onset and medical experience of the eight COVID-19 patients. Dates with possible exposure history to SARS-CoV-2 are marked in yellow and quarantine dates in pink. Real-time RT-PCR positive or negative for SARS-CoV-2 infection is represented by red or white star, respectively. Influenza A/B virus was not detected. The asterisks mark the dates of CT images shown in Figure S1 (for case 7) and Figure 4 (for case 8). Cities A and B are not in Hubei province.

Case 7 arrived from Wuhan on January 22 and was admitted into the hospital in quarantine with cough, high fever and lymphopenia on January 23 before the triaging period (Figure 3, Table S3). The chest CT progress with the ground glass shadow was consistent with the early imaging manifestation of viral pneumonia (Figure S1). Multiple rounds of nucleic acid analysis (at least 24 hours apart) before and after being transferred to the Section B failed to detect SARS-CoV-2 infection (Figure 3). The clinical features of the patient aggravated (Table S3). Subsequently, sputum of the patient was induced by 3% hypertonic saline nebulization and collected for RT-PCR analysis and the virus infection was confirmed (Figure 3). Of note, sputum-promoting operation was not routinely performed, as the effect of aerosol transmission of the virus indoor is of concern.

Before the epidemic alert, case 8 was admitted into the gastroenterology department due to retching for three weeks. During hospitalization (January 15– 22), other symptoms (shortness of breath, chest tightness), which were concealed before by retching, were revealed to the medical staff. He was diagnosed with polyserous effusions, constrictive pericarditis and lung infection and was discharged because of the alleviation of the above symptoms (Figure 3). Before triage, he visited the emergency department due to coughing. The number of leucocytes and lymphocytes was in normal range (Table 1). The chest CT images on January 26 showed bilateral pleural effusion and pericardial effusion (Figure 4A). On February 3, he was admitted to the ICU due to severe coughing and dyspnea with normal blood cell count, no recent suspicious epidemiological history, nor typical chest CT images of viral infection (Figure 4B, Table 1). Four days later, the chest CT images significantly changed and indicated possible viral infection. He was directed to the Section B (Figure 4C). However, the results from double nucleic acid analyses did not support for SARS-CoV-2 infection (Figure 3). Meanwhile, the symptoms (e.g. dyspnoea, chest tightness) were relieved with supportive treatments. However, the patient’s condition aggravated soon and he was sent into the emergency room. Since the chest CT images still indicated possible viral infection, multiple rounds of nucleic acid analysis were performed and confirmed SARS-CoV-2 infection (Figures 3 and 4D). Of note, both oropharyngeal and nasopharyngeal swabs were collected for nucleic acid analysis, but only the latter returned a positive result for SARS-CoV-2.

**Table 1.** Laboratory results of COVID-19 case 8.

|  | Jan 11 | Jan 15 | Jan 20 | Jan 26 | Feb 3 | Feb 7 | Feb 10 | Feb 15 | Feb 18 |
| --- | --- | --- | --- | --- | --- | --- | --- | --- | --- |
| White blood cell count, (3.5–9.5) × 10<sup>9</sup>/L | 5.4 | 5.9 | 4.2 | 4.7 | 4.6 | 5.8 | 4.8 | 5.5 | 5.3 |
| Hemoglobin, 115–150 g/L | 121.0 | 119.0 | 116.0 | 122.0 | 127.0 | 131.0 | 137.0 | 134.0 | 136.0 |
| Platelets, (125–350) × 10<sup>9</sup>/L | 268.0 | 210.0 | 137.0 | 179.0 | 171.0 | 122↓ | 109↓ | 77↓ | 82↓ |
| Neutrophils count, (1.8–6.3) × 10<sup>9</sup>/L | 3.9 | 4.6 | 3.2 | 3.1 | 3.2 | 4.5 | 3.9 | 4.6 | 4.2 |
| Neutrophils, 40–75% | 70.8 | 78.4↑ | 75.9↑ | 67.2 | 70.4 | 78.2↑ | 81.9↑ | 83.9↑ | 79.7↑ |
| Lymphocytes count, (1.1–3.2) × 10<sup>9</sup>/L | 1.1 | 0.8↓ | 0.6↓ | 1.1 | 1.2 | 0.8↓ | 0.5↓ | 0.6↓ | 0.6↓ |
| Lymphocytes, 20–50% | 19.9↓ | 13.5↓ | 13.3↓ | 23.6 | 25.3 | 13.9↓ | 10.2↓ | 10↓ | 11.4↓ |
| Albumin, 40–55 g/L | 35.2↓ | 32.4↓ | 28.3↓ | 33.9↓ | 29.5↓ | 28.9↓ | 27.6↓ | 28.6↓ | 32.6↓ |
| Alanine aminotransferase, 7–40 U/L | 395.9↑ | 532.8↑ | 368.7↑ | 137.8↑ | 237.7↑ | 163.9↑ | 227↑ | 147.5↑ | 88.2↑ |
| Aspartate aminotransferase, 13–35 U/L | 426.3↑ | 487.6↑ | 331↑ | 81.1↑ | 412.7↑ | 225↑ | 452.4↑ | 225.3↑ | 102.7↑ |
| Urea, 2.6–7.5 mmol/L | 10.1↑ | 11.6↑ | NA | NA | 14.2↑ | 8.3↑ | 9.5↑ | 7.5 | 6.9 |
| Creatinine, 41–111 μmol/L | 135.6↑ | 104.0 | NA | NA | 110.3 | 99.1 | 98.0 | 88.2 | 83.7 |
| Uric acid, 155–357 μmol/L | 340.6 | 339.3 | NA | NA | 393.9↑ | 313.1 | 259.9 | 126.9↓ | 107.9↓ |
| Lactate dehydrogenase, 120–250 U/L | 510↑ | 455↑ | 359↑ | 310↑ | 506↑ | NA | 522↑ | 385.9↑ | 362.3↑ |
| Creatine kinase, 40–200 U/L | 35.5↓ | 40.0 | 46.6 | 86.7 | 40.2 | NA | 80.8 | 135.5 | 64.8 |
| Creatine kinase isoenzyme, <24 U/L | 8.3 | 10.9 | 8.7 | 36.6↑ | 6.2 | NA | 13.4 | 19.0 | 20.6 |
| Myoglobin, <70 μg/L | 36.7 | 37.5 | 32.0 | 60.4 | 53.5 | NA | 113.7↑ | 209.6↑ | 137.8↑ |

NA: not available. ↑: above normal range. ↓: below normal range.

**Figure 4.**
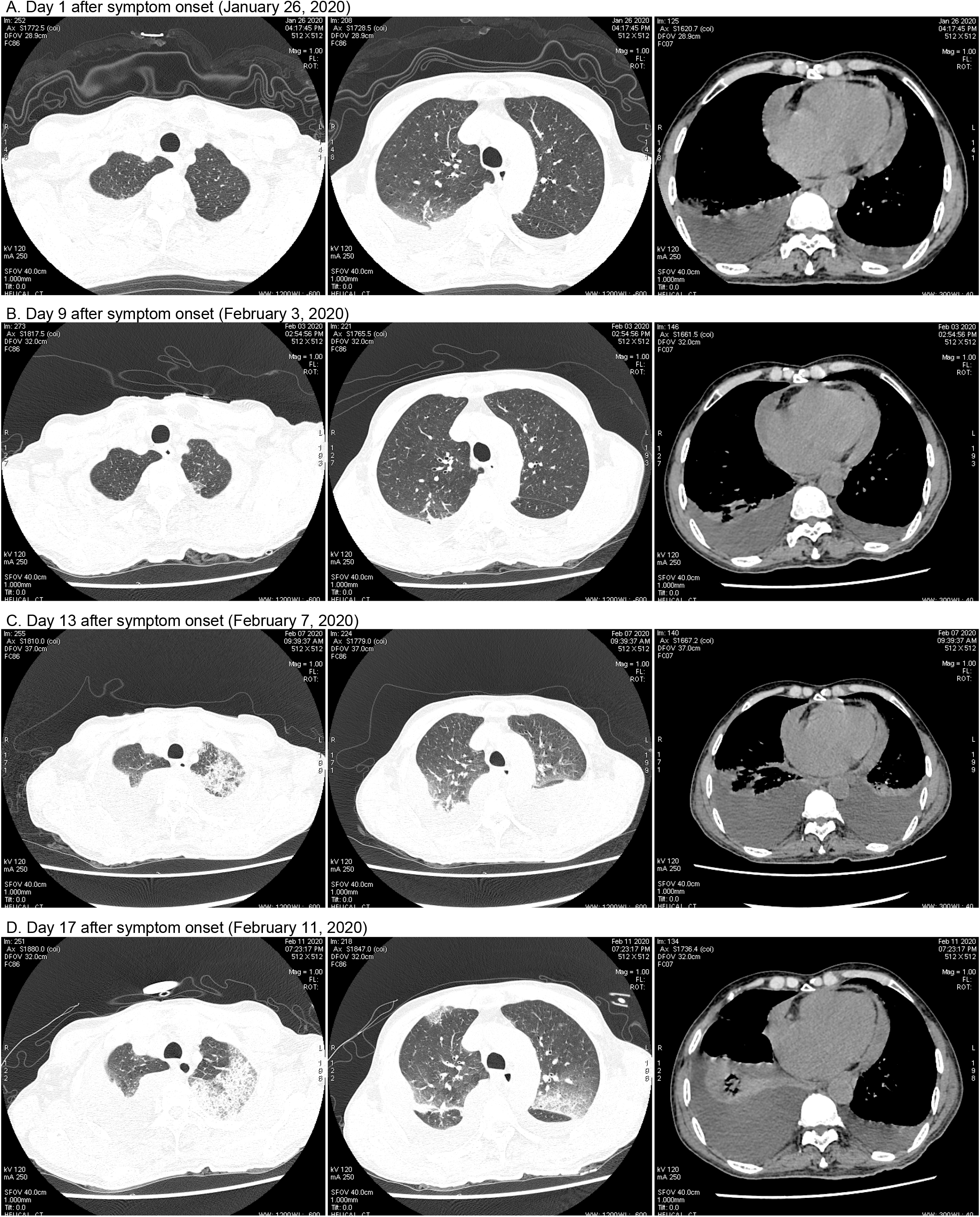
The course of chest CT images of case 8. (A) Images from January 26 show bilateral pleural effusion and pericardial effusion. (B) Images from February 3 show bilateral pleural effusion and pericardial effusion with a small area of ground-glass opacity in the left upper lung. (C) Images from February 7 show increased ground-glass opacity of the left upper lung with bilateral pleural effusion. A small area of new ground-glass opacity appeared in the right lung. (D) Images from February 11 show the further enlarged ground-glass opacity of the left upper lung. New ground-glass opacity is visible in the rest areas of lungs. The left pleural effusion decreased, and the right pleural effusion increased.

## DISCUSSION

Our retrospective study describes the clinical practice of triaging patients based on epidemiology in a local hospital of Changsha during the COVID-19 outbreak. The triage procedures last for 24 days covering the rapid spreading phase of SARS-CoV-2 in the city. Comparing to the situation of the Section B, patients with the virus infection were concentrated seven folds into the Section A, which was designated for rapid screening and quarantine. The first 10 days were shown to be critical in reducing the chance of spreading SARS-CoV-2. More than 85% of COVID-19 patients were identified during this period.

Rapid identification and isolation of COVID-19 patients was key to control the nosocomial infection, yet the pressure on Section B was still high. The virulence of SARS-CoV-2 seems to be weaker than SARS-CoV and MERS-CoV, but the ability of transmission among humans is stronger.^16^ During the 24 days of triage, roughly triple amounts of visits were received at the Section B as compared to the Section A. 247 individuals were suspected and received careful examination. More than 50% of them did multiple rounds of nucleic acid analysis at least 24 hours apart for signs of SARS-CoV-2. Eventually, eight COVID-19 cases were confirmed. Revisiting the patients diagnosed as negative for SARS-CoV-2 infection did not reveal that any COVID-19 patient was missed. Besides different course and severity of illness, delayed sharing of epidemiological history adds another layer of complexity to the diagnosis, underlining thorough diagnostic inquiry.

Most COVID-19 patients were identified as positive with one or two rounds of nucleic acid analysis in our study. Two cases (cases 7 and 8) seemed to be more complicated than usual. Case 7 arrived at the Section B with two negative nucleic acid reports already. The course of chest CT images, haematological features as well as previous Wuhan experience prompted three more rounds of nucleic acid analysis and finally confirmed the SARS-CoV-2 infection using the lower respiratory tract specimen. Case 8 had been enrolled into the hospital multiple times starting before the epidemic was alerted. Though being thoroughly examined, his comorbidities, no obvious recent epidemic history as well as the initial two negative results of nucleic acid analysis interfered the diagnosis. Accordingly, he was not in quarantine for treatment for roughly six days in the triage period. We only learned during the retrospective study that more than 200 people had returned to his town from Hubei province before January 25. Although this piece of information could not serve as concrete evidence for anything, it would have promoted the doctor to have second thoughts when looking at his case. After being finally confirmed as a COVID-19 case, overlapping patients along his track in the gastroenterology department and in the ICU were revisited. More than ten patients regarded as close contacts in the emergency department were quarantined for observation and diagnosis. Relevant medical stuff was submitted to CT/nucleic acid analysis. No one was found to be infected by SARS-CoV-2. Several additional measures were believed to contribute to this outcome: all patients were persuaded to actively wear masks during treatment in hospital area; disinfection of the hospital environment and medical stuff was implemented at least twice more frequently than usual during this period.

So far, no specific medication is available to cure COVID-19. Current clinical treatment is mainly supportive.^12^ The globalization of economy has facilitated the transmission of SARS-CoV-2 from one single city to six continents within very short time. When the daily reported new COVID-19 cases have been decreasing in China, they are continuously increasing in many other countries. On March 11, the WHO has characterized COVID-19 as a pandemic.^4^ The worldwide shortage of personal protective equipment has also been alerted. Triage — a medical practice that can be traced back to Napoleon’s time — has been integrated into daily practice in modern medical system.^17^ Yet, most clinicians, if not all, have no direct experience in the context of fighting a novel viral epidemic emergency. Our retrospective study of the triaging practice together with the diagnostic and clinical course of eight COVID-19 patients from 247 suspects will help other colleagues to control the transmission of the virus in the current COVID-19 outbreak.

## Data Availability

The data that support the findings of this study are available from the corresponding author on reasonable request. Participant data without names and identifiers will be made available after approval from the corresponding author and National Health Commission.

## SUPPLEMENTAL APPENDIX

Supplemental Appendix includes one figure (Figure S1) and three tables (Tables S1, S2 and S3) and can be found with this report online.

## ACKNOWLEDGMENTS

We thank Drs. Chengping Hu, Qiming Xiao (Department of Respiratory Medicine), Deming Tan (Department of Infectious Diseases), Xun Huang and Chunhui Li (Center for Healthcare-associated Infection Control) for their efforts in formulating the triage procedures; Drs. Yan Huang, Jun Quan and Fei Liu for facilitating data collection (Department of Infectious Diseases); Drs. Zhifei Zhan and Ge Zeng (Hunan Provincial Center for Disease Control and Prevention) for facilitating sample collection and analysis; Dr. Zhuohua Zhang (the Institute of Molecular Precision Medicine and Hunan Key Laboratory of Molecular Precision Medicine) for discussions and critical comments on the manuscript. The views expressed in this article are those of the authors and do not represent the official statement of Xiangya Hospital. The authors declare no competing interests. This work is supported by the National Natural Science Foundation (31700680, 31972886, 81803206), the Natural Science Foundation of Hunan Province (2018JJ2652, 2018JJ2667), the Scientific Research Project of Chinese Traditional Medicine Administration Bureau in Hunan Province (201806), the Research Projects from the Department of Science & Technology of Hunan province (2017RS3013, 2017XK2011, 2018DK2015, 2019SK1012, 2019RS1010), the Innovation-Driven Team Project from Central South University (2020CX016), China Postdoctoral Science Foundation (2018M632995), Xiangya Hospital Central South University Postdoctoral Foundation (to W. Zeng). None of the funders had any role in the study design, the collection/analysis/interpretation of data, the writing of the article and the decision to submit it for publication. The researchers confirm their independence from funders and sponsors.

